# Global Prevalence of Tobacco Smoking in People Living with Hepatitis C - Implications for Maximizing the Health Benefits from Antiviral Therapy: A Meta-Analysis

**DOI:** 10.1101/2025.09.23.25336463

**Authors:** Belaynew W Taye

## Abstract

Long-term antiviral treatment outcomes of hepatitis C infection may be affected by tobacco smoking. This study determined the prevalence of tobacco smoking in people living with hepatitis C virus (PLHCV) in low-and middle-income (LMICs) and high-income countries (HICs). We searched PubMed, EMBASE, PsycINFO, and ProQuest for studies published between January 1, 2008, and August 31, 2018. The quality of included studies was assessed using the Newcastle-Ottawa Scale. We performed meta-analysis using the Freeman-Tukey double arcsine transformation. We used Egger’s test to check for publication bias and performed meta-regression to identify individual-level sources of heterogeneity. The prevalence of tobacco smoking in PLHCV was 53.0%; it was 58% in LMICs and 52.0% in HICs. In subgroup analysis, the prevalence of tobacco smoking in PLHCV from clinic-based studies was 51% (95%CI 45%-57%) and it was 61% (95%CI 48%-73%) in community-based studies. In the multivariable meta-regression, study setting (coefficient=0.19, p<0.025) contributed significantly to the presence of heterogeneity between studies. Given the disproportionately high prevalence of tobacco smoking in PLHCV, addressing tobacco smoking in HCV treatment settings is recommended to maximise health benefits from antiviral therapy. That is particularly important in LMICs, where the burden of both tobacco smoking and HCV infection is growing.

## Introduction

Globally in 2016, hepatitis C virus (HCV) infection affected about 177.5 million (2.5%) people (Arnolfo et al., 2016) and many of the world’s top 10 high HCV burden countries are low– and middle-income countries (LMICs) (Thrift et al., 2017). Healthcare-associated HCV infection in LMICs remains an important risk factor contributing to a significant number of HCV infections annually (Mahmud et al., 2018; Mohlman et al., 2015).

The distribution of HCV infection prevalence by World Health Organization (WHO) regions varies by region. The highest rate of HCV infection was in the African Region (5.3%) followed by the Eastern Mediterranean (4.6%), and Western Pacific (3.9%) regions (Karoney & Siika, 2013). Globally in 2017, daily tobacco smoking prevalence was 15.2% (Peacock et al., 2018) with the majority (80%) of the world’s estimated 1.1 billion smokers living in LMICs(WHO, 2018). Tobacco smoking and HCV infection share similar risk factors and the prevalence of tobacco smoking might be high.

The presence of highly effective direct-acting antiviral treatment for HCV infection provides virologic cure to people with HCV infection and reversal of fibrosis and cirrhosis that are predominantly associated with development of hepatocellular carcinoma. Tobacco smoking in PLHCV is may have negative effects on the clinical progression of liver diseases after treatment with direct-acting antiviral drugs, which are associated with regression of liver fibrosis and significant reduction or prevention of hepatocellular carcinoma(Innes et al., 2018).

Understanding the burden of tobacco smoking in these patients wmay help plan to plan tobacco smoking cessation treatment during the course of treatment as well as during surveillance for screening for development of hepatocellular carcinoma.(Belaynew et al., 2018) This study aimed to estimate the prevalence of tobacco smoking in PLHCV in LMICs and HICs, and in the clinical and community-based samples. We discussed the implications for long-term health benefit of highly effective antiviral therapy for hepatitis C.

## Methods

The protocol was registered in PROSPERO (***CRD42019127774***) and the contents outlined in the PRISMA guidelines and flow diagram were adhered to for this meta-analysis (Moher et al., 2009).

### Literature search strategy

We searched PubMed, EMBASE, PsycINFO, and ProQuest databases for published observational studies on tobacco smoking and HCV infection in both HICs and LMICS. We used a combined medical subject heading (MeSH) and free-text terms for tobacco smoking, cigarette smoking, hepatitis C, prevalence, incidence, and epidemiology joined by the Boolean operators ‘AND’ and ‘OR’. The detailed search terms for each database are present in the additional file (**Supplementary file**).

### Eligibility Criteria

We included studies that assessed tobacco smoking in PLHCV in one of the LMICs or HICs (as classified by The World Bank Group)(Sumner, 2010) and published in English between January 1, 2008, and August 31, 2018. Grey literature was included by searching ProQuest database. To ensure consistency in measuring tobacco smoking (Edwards et al., 2013), we included studies with participants’ age ≥ 18 years and with HCV infection. We considered observational studies, including cross-sectional, comparative cross-sectional, survey, secondary data review, case-control, and retrospective or prospective cohort studies conducted in clinical or community settings.

We excluded narrative reviews, case reports, editorials and letters to editor, commentaries, viewpoints, systematic reviews, and meta-analyses as the required input data were lacking or were too small to be eligible for inclusion. Additionally, we excluded studies conducted in children, and with sample size below 40 participants.

All records retrieved from the four databases were exported to EndNote X8.2 *(EndNote. Clarivate Analytics, 2016)*. First, duplicate records were removed from the combined database. We screened for relevance of studies using title and abstract and records not relevant to tobacco smoking and HCV infection were excluded. We examined the full text of the remaining studies in detail to assess their eligibility. Finally, we assessed the methodological quality of studies that fulfilled the inclusion criteria and included in the meta-analysis.

### Study outcomes and measurements

The primary outcome of the study was tobacco smoking. We did not consider passive smoking, vaping, and the use of smokeless tobacco as tobacco smoking. Current tobacco use in included studies was defined as reported smoking of tobacco in the last five days before the survey irrespective of the frequency and number of cigarettes smoked or smoking tobacco on the day of data collection.(GTSS, 2009; Ryan et al., 2012) Some included studies defined tobacco smoking as 100 cigarettes in a lifetime, use of ≥ 1 cigarette per day in recent days, smoking ≥15 cigarettes per day, smoking ≥20 cigarettes per day or reported a history of tobacco smoking. Individual and study level variables that might be the source of heterogeneity were: study setting, year of study, The World Bank Group income classification, study quality score, study design, and WHO region.

### Risk of bias and quality assessment

Methodological quality and risk of bias in included studies were assessed using the NOS (Wells et al., 2012) instrument that is validated to evaluate the risk of bias in such studies. We directly used the NOS tools for cohort and case-control studies while we used the modified NOS to assess the methodological quality of cross-sectional studies.(Modesti et al., 2016) Using the guidelines for scoring utilizing this scale, we rated the quality of included articles and presented in a table.

### Data extraction

We extracted data on study variables relevant to the meta-analysis using standard electronic data extraction form created in Stata version 15.1 software. We included study identification, year of study, country of the study, World Bank income classification (LMIC or HIC), type of study population (general population, PLHCV, people living with human immunodeficiency virus (PLHIV), injecting drug users (IDU), dialysis patients, transplant patients), study setting (clinic, community), WHO region, sample size, and NOS quality score. Additionally, data on the tobacco smoking measurement method, the number of tobacco smokers, the number of PLHCV, and quality score were extracted and included. In situations where a study included adolescents below the age of 18 years, were extracted data by excluding participants < 18 years old from the results tables of these studies.

### Data synthesis and meta-analysis

We used the *metaprop* program of Stata 15.1 software (*StataCorp, 4905 Lakeway Drive, College Station, Tx USA*) to perform the meta-analysis. A random-effects meta-analysis was used to estimate the prevalence of tobacco smoking – due to the expected variation between regions and countries in their tobacco-smoking burden.

Freeman-Tukey double arcsine transformation (FTT) method was used to transform the proportions as the proportion of tobacco smoking for many included studies is closer to 1. Prevalence of tobacco smoking in PLHCV for sub-group and pooled estimates were performed by using DerSimonian–Liard non-iterative weighted estimates technique. Confidence intervals (CI) were calculated using the exact method. We used forest plots to display the prevalence of tobacco smoking in PLHCV and their confidence intervals.

We calculated the prevalence of tobacco smoking for each study by sub-groups based on income class, study setting, and WHO regions. Classification into LMICs and HICs was used because of the dynamic tobacco advertisement and marketing, leading to ever-changing prevalence and existence of significant variation of the context in these settings.

The presence of heterogeneity between included studies was tested using Cochrane’s Q statistic. The percentage of variation across studies that is due to heterogeneity, rather than chance, was measured by I^2^ statistic.

We performed a multivariable meta-regression analysis to identify the sources of heterogeneity and explore the effect of study-level independent variables on heterogeneity. The presence of publication bias was assessed by developing a funnel plot and checking for its symmetry. We also conducted Egger’s meta-bias test for the presence of small-study effects.

## Results

### Study selection

We identified 811 records from all databases searched and pooled them into one EndNote library. We removed 118 duplicate records. In the initial screening for relevance using title and abstracts, 593 studies were excluded for being unrelated to tobacco smoking in PLHCV. One hundred articles were relevant to this study, the full texts of these articles were examined for eligibility, and 42 articles fulfilled our inclusion criteria. The major reasons for exclusion were lack of data on both tobacco smoking and HCV infection (n=44), studies not published in English (n=2), very small sample size <40 (n= 5) and is a review and unable to extract tobacco smoking HCV infection data (n=7) (Figure 1).

**Figure 1.**
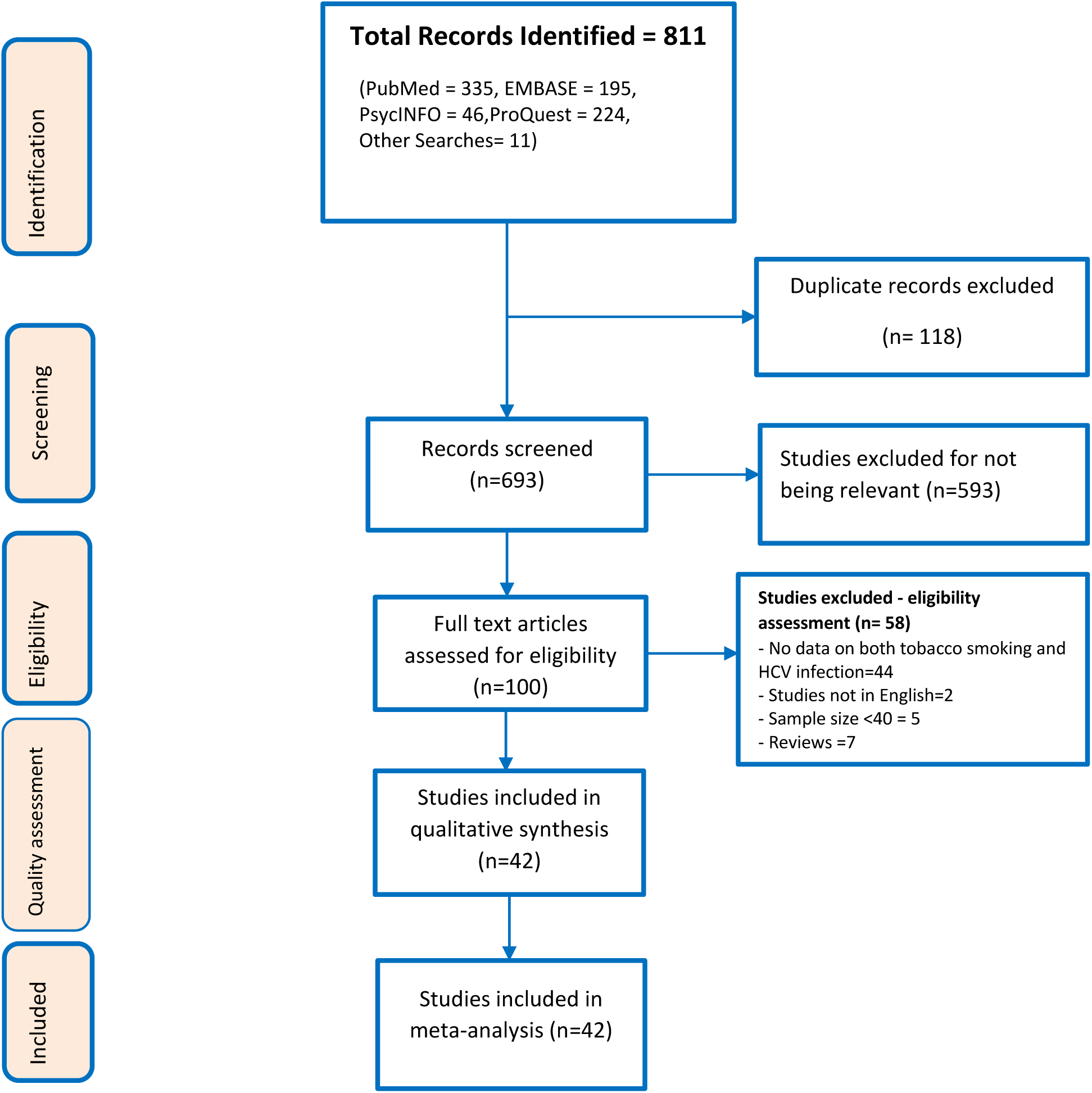
Preferred Reporting Items for Systematic Review and Meta-analysis flowchart for search and selection of articles reporting on tobacco smoking and hepatitis C virus infection in low, middle– and high-income countries.

### Characteristics of included studies

Eleven studies were from LMICs and 31 studies were conducted in HICs. (Supplementary file) Thirty-two studies were conducted in a clinical setting, while 10 were community-based studies. The majority (31out of 42) of studies were cross-sectional, and about a third (12 out of 42) were cohort studies. All included studies assessed HCV infection and tobacco smoking either as primary exposure and outcome or as part of the independent factors to measure other variables. Some assessed tobacco smoking as a comparison of PLHCV with HCV negative population and others compared tobacco smokers and non-smokers for treatment outcomes or health complications.

Overall, three-quarters (74.4%) of the studies were of high quality from the NOS assessment. All 12 cohort studies included in the meta-analysis had high NOS scores (six studies scored 9, and another six articles scored 8). Twenty-eight of the 38 cross-sectional studies had high (≥8) NOS scores (Supplementary file).

### Prevalence of tobacco smoking in people diagnosed with hepatitis C

In all regions of the world, the pooled prevalence of tobacco smoking in PLHCV was 53.0% (95% CI 47%-60%).

### Low-and middle-income and high-income countries

Twelve studies were from LMICs, and the prevalence of tobacco smoking in PLHCV was 58% (95%CI 37%-78%). In LMICs, the highest prevalence observed was 100% and the lowest was 3%. In half of the studies, the prevalence of tobacco smoking in PLHCV living in LMICs was higher than 50%. In HICs, the prevalence of tobacco smoking in PLHCV was 52.0% (95%CI 45%-59%). The highest prevalence was 88% while the smallest was 13%. There was high and significant heterogeneity between LMICs and HICs and prevalence of tobacco smoking among these sub-groups was not pooled (Figure 2).

**Figure 2.**
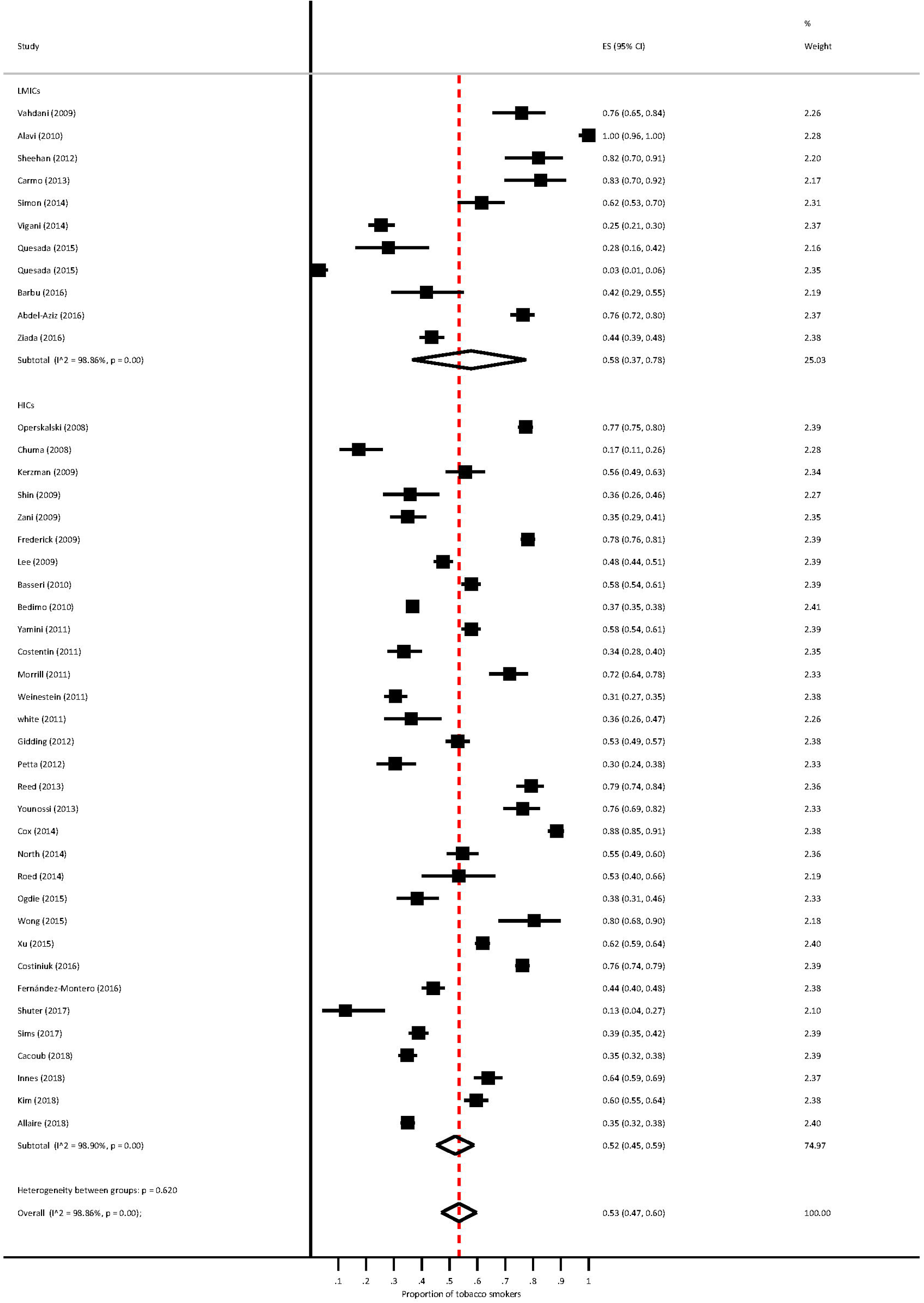
Prevalence of tobacco smoking in people living with hepatitis C in low, middle-income, and high-income countries, 2008-2018. It presents a forest plot of the random effects meta-analysis of pooled prevalence of tobacco smoking in people living with hepatitis C virus in low-and middle-income and high-income countries. The boxes show effect estimate (prevalence) in individual studies, the bold lines represent confidence intervals, and diamond represents pooled prevalence in LMICs, HICs and overall estimate.

### Clinical and community-based samples

Thirty-two studies were conducted in a clinical setting. The heterogeneity test between the clinical and community-based studies for the hypothesis was not significant, and the groups were homogeneous. The prevalence of tobacco smoking in PLHCV in clinic-based studies was 51% (95%CI 45%-57%).

The pooled prevalence of tobacco smoking in PLHCV from11 community-based studies was 61% (95%CI 48%-73%). The lowest tobacco smoking prevalence in PLHCV was 3%, and the highest was 88%.

An interesting observation in clinical studies is that while there was heterogeneity between studies until 2014, the prevalence of tobacco smoking showed a decline and was to the left of the central line. On the other hand, community-based studies showed a higher prevalence of tobacco smoking in PLHCV during 2008 to 2018 (Figure 3).

**Figure 3.**
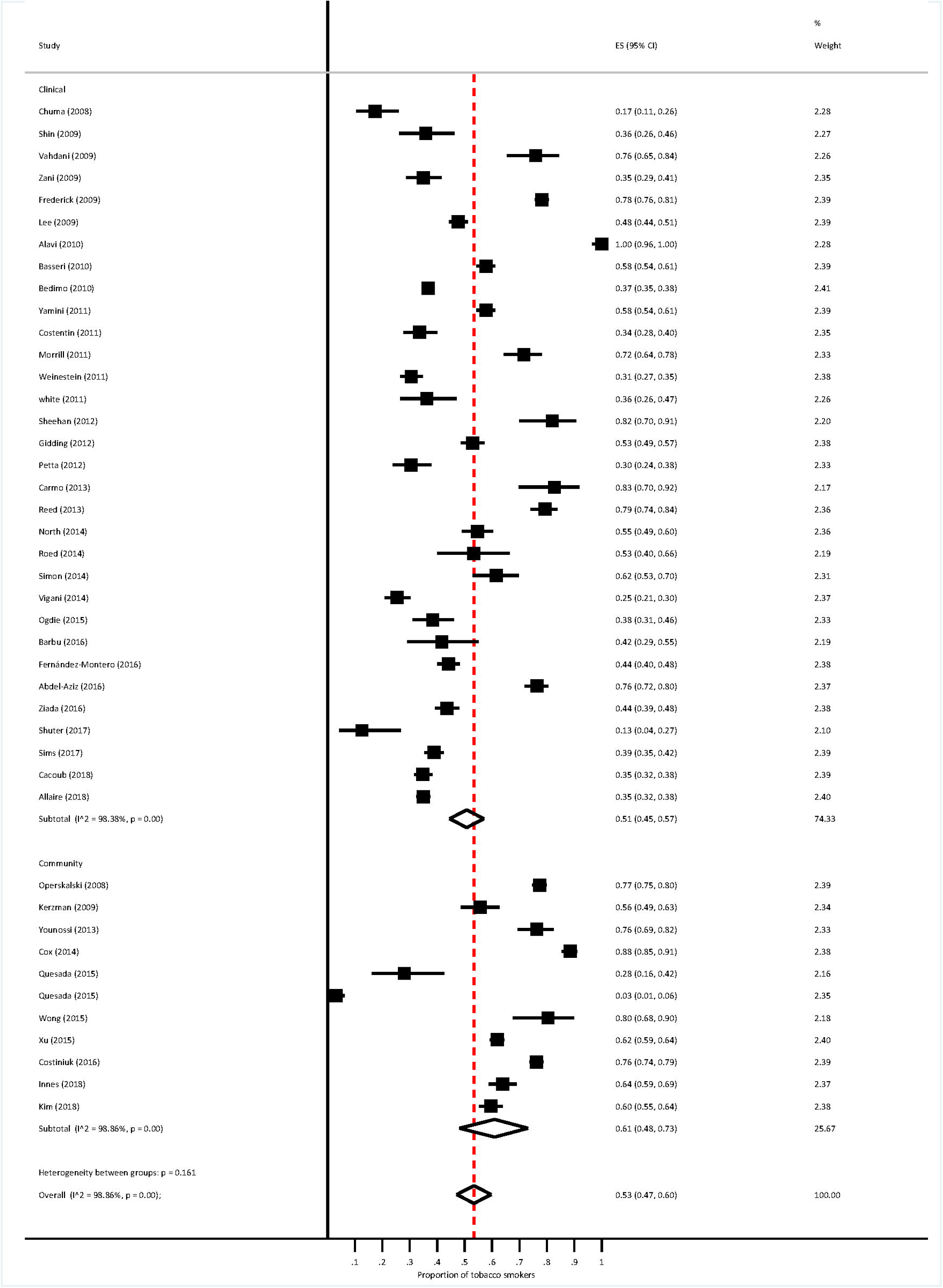
Prevalence of tobacco smoking in people living with hepatitis C virus in clinical and community-based samples, 2008 – 2018. It presents a forest plot of the random effects subgroup meta-analysis for pooled prevalence of tobacco smoking in people living with hepatitis C virus in clinical and community-based samples. The boxes show effect estimate (prevalence) in individual studies, the bold lines represent confidence intervals, and the diamond shapes represent pooled prevalence in clinical, community and overall samples.

### Tobacco smoking prevalence by WHO regions

There were studies in all WHO regions, but WPRO (1 study) and EMRO (2 studies) had very few included studies. The highest prevalence of tobacco smoking in PLHCV was 94% (95%CI 90%-97%) in the EMRO followed by AMRO, 60 % (95%CI 51%-68%), WPRO (57%), AFRO (37%), and SEARO (35%).

### Publication Bias

A funnel plot was produced to assess for the presence of publication bias in the review. The funnel plot is symmetric, showing the absence of small-study effects. Egger’s meta-bias test showed no small-study effects as there was not enough evidence to reject the null hypothesis of “no small-study effects” with a bias = 1.45 (95%CI –1.01-3.90), p-value= 0.242 (Figure 4).

**Figure 4.**
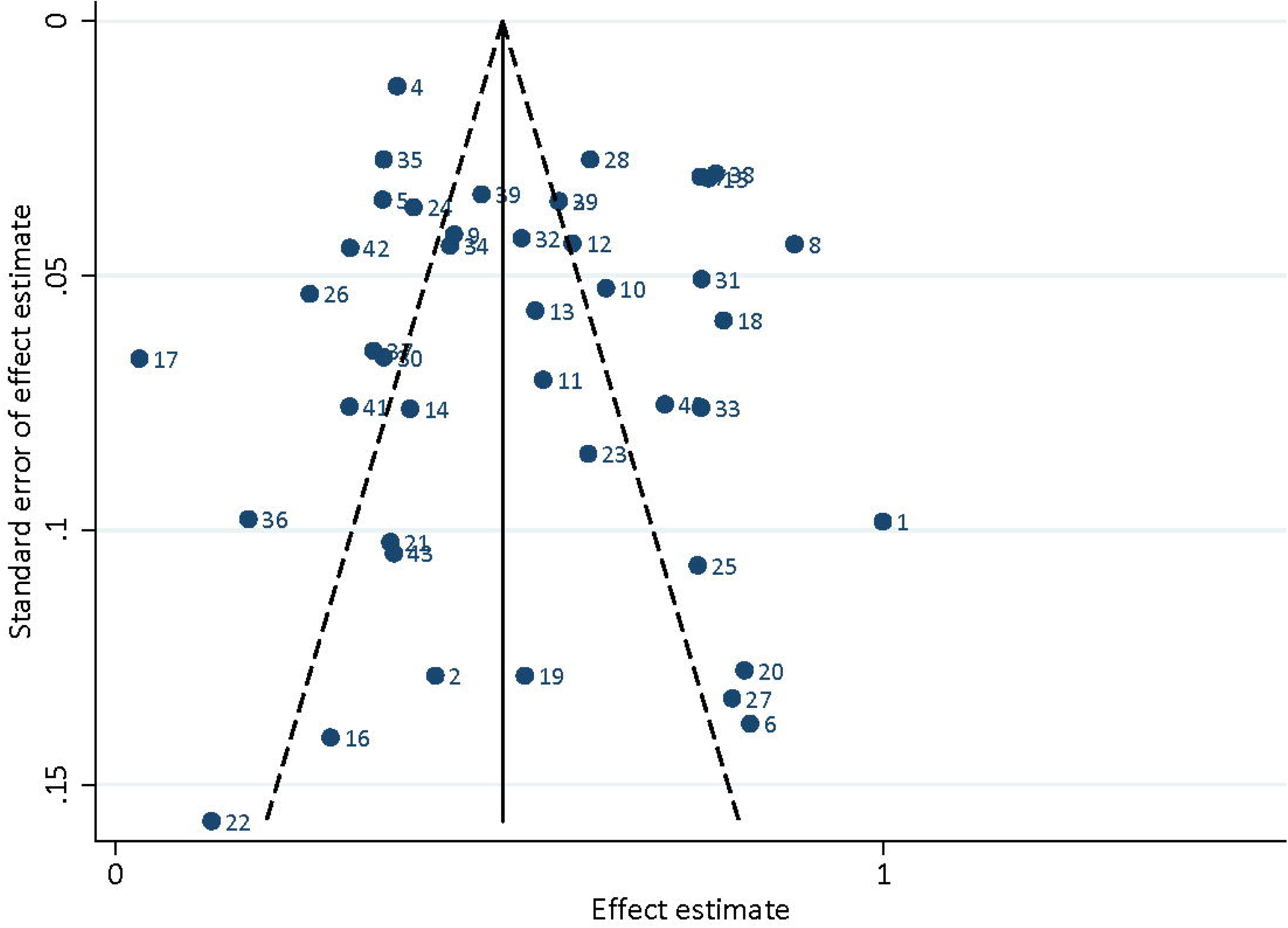
Funnel plot to assess publication bias of studies assessing tobacco smoking and HCV infection. Figure 5 represents a funnel plot that shows effect estimate against its standard error. Bold dots and numbers represent individual studies. The symmetric funnel reflects the absence of publication bias and small study effects.

### Factors for heterogeneity between studies (meta-regression)

In the univariate analysis, none of the variables were significant contributors to heterogeneity, but in the multivariable meta-regression, study setting (p<0.025) significantly contributed to heterogeneity. However, WHO region (p=0.280), income class (p=0.344), study year (p=0.069), study design (p=0.15) and NOS score (p>0.05) were not significantly associated with presence of heterogeneity (Table 1).

**Table 1.** Meta-regression analysis for sources of heterogeneity on studies assessing tobacco smoking and HCV infection.

| Character | Coefficient | Std.error | t | P>\|t\| | 95% Confidence interval |  |
| --- | --- | --- | --- | --- | --- | --- |
| WHO region | -0.03 | 0.03 | -1.10 | 0.280 | -0.094 | 0.028 |
| Income class | -0.08 | 0.08 | -0.96 | 0.344 | -0.246 | 0.088 |
| Year | -0.02 | 0.01 | -1.87 | 0.069 | -0.048 | 0.002 |
| Study design | 0.06 | 0.04 | 1.46 | 0.152 | -0.022 | 0.137 |
| NOS score | -0.04 | 0.03 | -1.31 | 0.198 | -0.110 | 0.024 |
| Setting | 0.19 | 0.08 | 2.33 | 0.025 | 0.025 | 0.361 |
| Constant | 47.07 | 24.74 | 1.90 | 0.065 | -3.098 | 97.233 |

## Discussion

We performed a systematic review and meta-analysis to estimate the prevalence of tobacco smoking in PLHCV in LMICs and HICs. A total of 43 tobacco smoking and HCV infection-related data were extracted from 42 studies (one study(Quesada et al., 2015) had included assessments from eight countries, two of which are eligible to be included in the analysis).

The studies assessed tobacco smoking and HCV infection directly or indirectly as part of the measurement of treatment response of DAAs, comparative survey of PLHCV and HCV negative persons, renal or liver transplant patients. Tobacco smoking prevalence studies in PLHCV are lacking, and data on the number of PLHCV and the number of tobacco smokers from observational studies that measured tobacco smoking and HCV infection were extracted to calculate prevalence. Moreover, the burden of tobacco smoking in PLHCV is unknown in LMICs or HICs and the WHO regions of the world. Obtaining data from PLHCV on tobacco smoking and determining the prevalence, therefore, provides a valuable alternative to know the burden of tobacco smoking among special populations.

Over half of the PLHCV globally (53%) are exposed to tobacco smoking compared to 15.2% in the general population,(Peacock et al., 2018) a prevalence that is more than three times higher. The very high burden of tobacco smoking may be due to the common underlying risk factors shared between tobacco smoking and HCV infection including illicit substance use, unsafe IDU, and depression and anxiety(Kim et al., 2018; Shuter et al., 2017) which leads to the high concentration of tobacco smokers in people diagnosed with HCV. Moreover, many beliefs and socio-economic conditions of PLHCV may prompt them to start smoking.

In LMICs, tobacco smoking prevalence in PLHCV was higher than tobacco smoking prevalence in HICs. The higher prevalence of tobacco smoking in LMICs could be a reflection of the global pattern of tobacco smoking in the general population, where the rate of tobacco smoking is declining in HICs but not the case in LMICs (Ng et al., 2014). Furthermore, there is a shift in the marketing of tobacco products towards LMICs in Africa, Asia, and Eastern Europe (WHO, 2013) in search of the tobacco market. Other reasons may be rapid population growth, lack of proper tobacco control measures and poor access to smoking cessation interventions also play a vital role in the changing epidemiology of tobacco smoking in LMICs (WHO, 2013).

In community-based populations, tobacco smoking prevalence was higher than the prevalence among clinic-based populations. This could be because clinical populations have better health awareness and refrain from smoking tobacco. Additionally, clinic-based populations have morbidities from HCV infection or other diseases, and current smoking rates could be lower due to cessation interventions or quitting earlier.

We observed a decreasing prevalence of tobacco smoking in clinic-based populations from 2015 onwards but increasing in the community-based studies. Population-based surveys that include a wide range of PLHCV from LMICs and HICs in the community and clinical settings are required to reflect the real picture of risk factors for tobacco smoking in these populations.

The higher burden of tobacco smoking in PLHCV poses a critical challenge to adherence to and treatment success of DAAs for HCV infection (El-Zayadi, 2006; Fan et al., 2015; Fujita et al., 2006), particularly in LMICs unless proper tobacco smoking cessation and harm reduction strategies are implemented. A cross-sectional study of HCV positive blood donors in Israel (Kerzman et al., 2009) reported that smoking was found to be a significant predictor of non-compliance with recommendations to seek medical counselling from general practitioners or liver specialists who may apply to PLHCV. Therefore, PLHCV on DAA therapy could face challenges in adherence to the clinical and behavioural recommendations essential to treat HCV infection aimed at altering the natural history of chronic HCV infection with long-term impact on liver and other systems. Belaynew et’ al also identified a potential adverse effect of tobacco smoking on SVR among interferon-based antivirals for HCV infection treatment although there was no evidence about DAAs.(Belaynew et al., 2018)

Study setting was a significant independent determinant of heterogeneity between studies. The reason for this is not clear but it might be due to the differentials in risks behaviours among clinical and community-based populations. Clinic-based populations, for example, included patients with several morbidities like diabetes, renal or liver transplant that might have undergone a series of counselling sessions to stop smoking, while patients from community-based population may not have received such counselling. Additionally, patients from clinic-based studies might have utterly different health behaviours that led to health seeking compared to the general population. This implies that the prevalence of tobacco smoking shall be interpreted separately using the sub-group analysis.

### Implications for health benefits gained from direc-acting antivirals

Viral eradication with antiviral treatment provides reversal of liver inflammation and fibrosis and prevents hepatocellular carcinoma, cardiovascular diseases, diabetes and renal diseases(Cacoub et al., 2018). High burden of tobacco smoking in people with hepatitis C undermines the therapeutic benefits from the highly efficacious direct-acting anti-viral therapy. Reversal or regression of liver fibrosis after direct-acting antiviral treatment may not be possible due to the effect of tobacco smoking on the liver that can accelerate the clinical progression of chronic liver disease. Persons with hepatitis C who smoke tobacco may present with advanced liver disease due to interaction between the two risk factors. Tobacco contains chemicals that result in several chronic diseases including cirrhosis and hepatocellular carcinoma. These are also caused by HCV infection (Glantz & Gonzalez, 2012) and as a result, tobacco smoking substantially increases morbidity and mortality in these populations. Tobacco smoking and HCV infection interact to cause liver fibrosis and hepatocellular carcinoma (HCC)(Chuang et al., 2010), chronic respiratory diseases, cardiovascular diseases, diabetes mellitus, and chronic renal diseases (Belaynew et al., 2018) Moreover, tobacco smoking in PLHCV affects adherence (Kerzman et al., 2009), treatment response, and progression of HCV infection. Concomitant harm reduction treatment in anti-HCV treatment settings provides additional opportunities to prevent chronic diseases including diseases of the liver, cardiovascular system, and the kidney(Fan et al., 2015; Glantz & Gonzalez, 2012). Tobacco harm reduction in PLHCV improves the quality of life of these patients after completion of antiviral therapy (Yamini et al., 2011).

This meta-analysis estimated the global burden of tobacco smoking in PLHCV and described by income classification (very important driver of both tobacco smoking and HCV infection), study setting and WHO regions. This provides an insight to planning effective interventions. Data on tobacco smoking and HCV infection were partly extracted from clinic-based observational studies that may not replace community-based surveys, which are random samples of the general population aimed to determine prevalence.

In conclusion, we found tobacco smoking in PLHCV is disproportionately higher than the prevalence in the general population, with higher rates in PLHCV from LMICs and community-based populations. The higher rate of tobacco smoking in PLHCV in LMICs could be due to the marketing of tobacco products towards LMICs such as in Africa, Asia, and Eastern Europe.

Integrated tobacco smoking cessation and harm reduction interventions at HCV treatment settings are recommended to maximise the health benefits from anti-HCV therapy and improve the quality of life of patients. Population-based studies covering different socio-economic, geographic, and clinical settings are recommended to identify the risk factors for tobacco smoking in PLHCV.

## Data Availability

The data used in to produce this study are included in the article or its supplementary files.

## Acknowledgements

Not applicable.

## References

1. Arnolfo, Petruzziello, Samantha, Marigliano, Giovanna, Loquercio, et al. (2016). Global epidemiology of hepatitis C virus infection: an update of the distribution and circulation of hepatitis C virus genotypes. World J Gastroenterol (34), 7824–7840.

2. Belaynew, T., Patricia, V., Malcolm, B., Linda, S., Charles, G., Coral, G., et al. (2018). Tobacco Smoking and Hepatitis C Virus Infection: Burden, Health Effects and Opportunities for Treatment of Both The University of Queensland, Faculty of Medicine.

3. Cacoub, P., Nahon, P., Layese, R., Blaise, L., Desbois, A. C., Bourcier, V., et al. (2018). Prognostic value of viral eradication for major adverse cardiovascular events in hepatitis C cirrhotic patients. Am Heart J, 198, 4–17. 10.1016/j.ahj.2017.10.024

4. Chuang, S. C., Lee, Y. C., Hashibe, M., Dai, M., Zheng, T., & Boffetta, P. (2010). Interaction between cigarette smoking and hepatitis B and C virus infection on the risk of liver cancer: a meta-analysis. Cancer Epidemiol Biomarkers Prev, 19(5), 1261–1268. 10.1158/1055-9965.Epi-09-1297

5. Edwards, R., Carter, K., Peace, J., & Blakely, T. (2013). An examination of smoking initiation rates by age: results from a large longitudinal study in New Zealand. Aust N Z J Public Health, 37(6), 516–519. https://onlinelibrary.wiley.com/doi/pdf/10.1111/1753-6405.12105, https://onlinelibrary.wiley.com/doi/pdfdirect/10.1111/1753-6405.12105?download=true

6. El-Zayadi, A. R. (2006). Heavy smoking and liver. World J Gastroenterol, 12(38), 6098–6101. https://www.ncbi.nlm.nih.gov/pubmed/17036378, https://www.ncbi.nlm.nih.gov/pmc/articles/PMC4088100/pdf/WJG-12-6098.pdf

7. Fan, J. Y., Huang, T. J., Jane, S. W., & Chen, M. Y. (2015). Prevention of Liver Cancer Through the Early Detection of Risk-related Behavior Among Hepatitis B or C Carriers. Cancer Nurs, 38(3), 169–176. 10.1097/ncc.0000000000000153

8. Fujita, Y., Shibata, A., Ogimoto, I., Kurozawa, Y., Nose, T., Yoshimura, T., et al. (2006). The effect of interaction between hepatitis C virus and cigarette smoking on the risk of hepatocellular carcinoma. Br J Cancer, 94(5), 737–739. 10.1038/sj.bjc.6602981

9. Glantz, S., & Gonzalez, M. (2012). Effective tobacco control is key to rapid progress in reduction of non-communicable diseases. The Lancet, 379(9822), 1269–1271. 10.1016/S0140-6736(11)60615-6

10. GTSS, Global Tobacco Surveillance System. (2009). Global Adult Tobacco Survey (GATS). Indicator Guidelines: Definition and Syntax. 2009 Retrieved from http://www.who.int/tobacco/surveillance/en_tfi_gats_indicator_guidelines.pdf

11. Innes, H., McAuley, A., Alavi, M., Valerio, H., Goldberg, D., & Hutchinson, S. J. (2018). The contribution of health risk behaviors to excess mortality in American adults with chronic hepatitis C: A population cohort-study [Article]. Hepatology, 67(1), 97–107. 10.1002/hep.29419

12. Karoney, M. J., & Siika, A. M. (2013). Hepatitis C virus (HCV) infection in Africa: a review. The Pan African medical journal, 14, 44–44. 10.11604/pamj.2013.14.44.2199

13. Kerzman, H., Green, M. S., & Shinar, E. (2009). Predictors for non-compliance of notified hepatitis C virus-positive blood donors with recommendation to seek medical counselling. Vox Sang, 96(1), 20–28. 10.1111/j.1423-0410.2008.01111.x

14. Kim, R. S., Weinberger, A. H., Chander, G., Sulkowski, M. S., Norton, B., & Shuter, J. (2018). Cigarette Smoking in Persons Living with Hepatitis C: The National Health and Nutrition Examination Survey (NHANES), 1999-2014. Am J Med, 131(6), 669–675. 10.1016/j.amjmed.2018.01.011

15. Mahmud, S., Kouyoumjian, S. P., Al Kanaani, Z., Chemaitelly, H., & Abu-Raddad, L. J. (2018). Individual-level key associations and modes of exposure for hepatitis C virus infection in the Middle East and North Africa: a systematic synthesis. Ann Epidemiol, 28(7), 452–461. 10.1016/j.annepidem.2018.03.007

16. Modesti, P. A., Reboldi, G., Cappuccio, F. P., Agyemang, C., Remuzzi, G., Rapi, S., et al. (2016). Panethnic Differences in Blood Pressure in Europe: A Systematic Review and Meta-Analysis. PLoS One, 11(1), e0147601. 10.1371/journal.pone.0147601

17. Moher, D., Liberati, A., Tetzlaff, J., Altman, D. G., & Group, P. (2009). Preferred reporting items for systematic reviews and meta-analyses: the PRISMA statement. J Clin Epidemiol, 62(10), 1006–1012. 10.1016/j.jclinepi.2009.06.005

18. Mohlman, M. K., Saleh, D. A., Ezzat, S., Abdel-Hamid, M., Korba, B., Shetty, K., et al. (2015). Viral transmission risk factors in an Egyptian population with high hepatitis C prevalence. BMC Public Health, 15, 1030. 10.1186/s12889-015-2369-y

19. Ng, M., Freeman, M. K., Fleming, T. D., Robinson, M., Dwyer-Lindgren, L., Thomson, B., et al. (2014). Smoking prevalence and cigarette consumption in 187 countries, 1980-2012. JAMA, 311(2), 183–192. 10.1001/jama.2013.284692

20. Peacock, A., Leung, J., Larney, S., Colledge, S., Hickman, M., Rehm, J., et al. (2018). Global statistics on alcohol, tobacco and illicit drug use: 2017 status report. Addiction, 113(10), 1905–1926. 10.1111/add.14234

21. Quesada, P., Whitby, D., Benavente, Y., Miley, W., Labo, N., Chichareon, S., et al. (2015). Hepatitis C virus seroprevalence in the general female population from 8 countries. J Clin Virol, 68, 89–93. 10.1016/j.jcv.2015.05.005

22. Ryan, H., Trosclair, A., & Gfroerer, J. (2012). Adult current smoking: differences in definitions and prevalence estimates--NHIS and NSDUH, 2008. J Environ Public Health, 2012, 918368. 10.1155/2012/918368

23. Shuter, J., Litwin, A. H., Sulkowski, M. S., Feinstein, A., Bursky-Tammam, A., Maslak, S., et al. (2017). Cigarette Smoking Behaviors and Beliefs in Persons Living With Hepatitis C [Article]. Nicotine Tob Res, 19(7), 836–844. 10.1093/ntr/ntw212

24. Sumner, A. (2010). Global Poverty and the New Bottom Billion: What if Three-quarters of the World’s Poor Live in Middle-income Countries? IDS Working Papers, 2010(349), 01–43. 10.1111/j.2040-0209.2010.00349_2.x

25. Thrift, A. P., El-Serag, H. B., & Kanwal, F. (2017). Global epidemiology and burden of HCV infection and HCV-related disease. Nat Rev Gastroenterol Hepatol, 14(2), 122–132. 10.1038/nrgastro.2016.176

26. Wells, G., Shea, B., O’Connell, D., Peterson, J., Welch, V., & Losos, M. (2012). The Newcastle-Ottawa Scale (NOS) for assessing the quality of nonrandomised studies in meta-analyses. Ottawa, Ontario, Canada: Ottawa Hospital Research Institute; 2013. In.

27. WHO, World Health Organization. (2013). Tobacco Free Initiative. Retrieved from Accessed online at: http://www.who.int/tobacco/mpower/en/

28. WHO, World Health Organization. (2018). *Tobacco* http://www.who.int/news-room/fact-sheets/detail/tobacco

29. Yamini, D., Basseri, B., Chee, G. M., Arakelyan, A., Enayati, P., Tran, T. T., et al. (2011). Tobacco and other factors have a negative impact on quality of life in hepatitis C patients [Article]. J Viral Hepat, 18(10), 714–720. 10.1111/j.1365-2893.2010.01361.x

