## Supplementary File for "Global Prevalence of Tobacco Smoking in People Living with Hepatitis C - Implications for Maximizing the Health Benefits from Antiviral Therapy: A Meta-Analysis"

### Supplementary material

#### A. Search strategy used in meta-analysis of tobacco smoking prevalence in people living with hepatitis C virus

**PubMed**

Epidemiolog* OR "Epidemiology"[Mesh] OR Prevalence OR "Prevalence"[Mesh] OR Incidence OR "Incidence"[Mesh] AND "Smoking"[Mesh:NoExp] OR "Tobacco Smoking"[Mesh] OR cigar*[tiab] OR tobacco[tiab] AND "hepatitis C" OR hepacivirus OR "Hepacivirus"[Mesh:NoExp] OR "Hepatitis C"[Mesh] OR HCV.

**EMBASE**

prevalence OR 'epidemiology' OR incidence) AND ('hepacivirus':ab,ti OR 'hepatitis c':ab,ti OR 'hepatitis c virus':ab,ti OR 'chronic hepatitis c':ab,ti) AND ('cigarette smoking':ab,ti OR 'smoking':ab,ti OR 'cigar smoking':ab,ti) AND human AND adult AND ('case control study' OR 'cohort analysis' OR 'comparative study' OR 'cross-sectional study' OR 'interview' OR 'longitudinal study' OR 'medical record review' OR 'multicenter study' OR 'observational study' OR 'prospective study' OR 'questionnaire' OR 'retrospective study' while to retrieve records from

**PsycINFO**

prevalence OR Any Field: Epidemiology AND Any Field: tobacco smoking OR Any Field: smoking AND Any Field: hepatitis C virus infection OR Any Field: Hepatitis C.

**ProQuest**

MAINSUBJECT.EXACT("Cigarettes") OR MAINSUBJECT.EXACT("Cigars") OR ("tobacco smoking")) AND (("hepatitis c virus") OR ("chronic hepatitis C")) AND (prevalence OR incidence OR Epidemiology).

**B**

Supplementary Table 1. Characteristics of included studies that assessed tobacco smoking in people living with hepatitis C virus

| Study ID | Study period | Country | WHO Region | Setting | Age | Population type | Income class (Worldbank) | Study Design | Sample size | Number of PLHCV | Tobacco smoking measurement | Number of Smokers | Risk of bias score |
|---|---|---|---|---|---|---|---|---|---|---|---|---|---|
| Abdel-Aziz, 2016 | 2013 - 2014 | Egypt | Africa | Clinic | 18+ | PLHCV/HCV- | LMICs | Case-control | 778 | 389 | Not stated | 297 | 9 |
| Alavi, 2010 | 2002 - 2006 | Iran | Middle East | Clinic | 24.8±6.2 | IVDU | LMICs | Cross-sectional | 333 | 103 | Current use | 103 | 7 |
| Allaire, 2018 | 2006 - 2012 | France | EURO | Clinic | >18 | PLHCV/HBV | HIC | Cohort | 1671 | 1354 | Current use | 473 | 9 |
| Barbu, 2016 | 2015 - 2016 | Romania | Europe | Clinic | 18+ | PLHCV | LMICs | Cross-sectional | 120 | 60 | Current use | 25 | 6 |
| Basseri, 2010 | 1998 - 2007 | USA | North America | Clinic | 18+ | PLHCV | HICs | Cross-sectional | 800 | 800 | Current use | 462 | 8 |
| Bedimo, 2010 | 1996 - 2004 | USA | North America | Clinic | 46.2±10.2 | PLHIV | HICs | Cohort | 19424 | 6136 | Past/current use | 2251 | 8 |
| Cacoub, 2018 | 2006-2012 | France | Europe | Clinic | 18+ | PLHCV | HICs | Cohort | 878 | 813 | Current use | 283 | 8 |
| Carmo, 2013 | 2005 - 2007 | Brazil | Latin America | Clinic | 18+ | Mental illness | LMICs | Cross-sectional | 2087 | 52 | Current use | 43 | 9 |
| Chuma, 2008 | 1987 - 2002 | Japan | SEARO | clinic | 50.5±11.5 | Cirrhosis | HIC | Cohort | 104 | 104 | ≥20 pack-yrs. | 18 | 9 |
| Costentin, 2011 | 2004-2006 | France | EURO | Clinic | 45±11 | PLHCV | HIC | Cross-sectional | 238 | 238 | >15cig/day | 80 | 8 |
| Costiniuk, 2016 | 2003 - 2014 | Canada | North America | Community | 16+ | PLHCV/HIV | HICs | Cohort | 1072 | 1072 | Current use | 817 | 8 |
| Cox, 2014 | 2003-2013 | Canada | AMRO | Clinic | 44(38 - 49) | PLHCV/HIV | HIC | Cohort | 521 | 521 | Current use | 461 | 9 |
| Fernández-M, 2016 | 2004 -2014 | Spain | Europe | Clinic | 42.7 | PLHIV | HICs | Cohort | 1136 | 569 | Current use | 251 | 9 |
| Frederick, 2009 | 1994-2002 | USA | AMRO | Clinic | 18+ | PLHCV/HIV | HIC | Cross-sectional | 3636 | 1115 | Current use | 872 | 10 |
| Gidding, 2012 | 2008 - 2009 | Australia | Australia | Clinic | 46(median) | PLHCV | HICs | Cohort | 550 | 550 | Current use | 291 | 8 |
| Innes, 2018 | 1999-2010 | USA | North America | Clinic | 20+ | PLHCV/HIV | HICs | Cohort | 27468 | 363 | Current use/>100 cig/life | 232 | 9 |
| Kerzman, 2009 | 2001–2002 | Israel | Middle East | Community | 31·2 ± 10·8 | PLHCV blood donors | LMICs | Cross-sectional | 201 | 201 | Current use | 112 | 8 |
| Kim, 2018 | 1999-2014 | USA | North America | Community | ≥20 | PLHCV | HICs | Cross-sectional | 39472 | 524 | Current use/ past 5 days | 312 | 9 |
| Lee, 2009 | 1999 - 2007 | USA | AMRO | Clinic | 49.3±8.5 | CLD | HIC | cohort | 2260 | 864 | Active smoker | 412 | 9 |
| Morrill, 2011 | 2000 - 2008 | USA | AMRO | Clinic | 18+ | PLHCV | HIC | Cross-sectional | 176 | 176 | Not stated | 126 | 6 |
| North, 2014 | 2014 | USA | North America | Clinic | 52.6±7.5 | PLHCV | HICs | Cross-sectional | 309 | 309 | Current use | 169 | 6 |
| Ogdie, 2015 | 2008-2012 | USA | North America | Clinic | 18-80 | PLHCV/HIV | HICs | Cross-sectional | 202 | 172 | Current use | 66 | 8 |
| Operskalski, 2008 | 1994-2002 | USA | North America | Community | >35 | PLHCV women | HICs | Cross-sectional | 1049 | 1049 | Current use | 811 | 8 |

| Study ID | Study period | Country | WHO Region | Setting | Age | Population type | Income class (Worldbank) | Study Design | Sample size | Number of PLHCV | Tobacco smoking measurement | Number of Smokers | Risk of bias score |
|---|---|---|---|---|---|---|---|---|---|---|---|---|---|
| Petta, 2012 | 2012 | Italy | EURO | Clinic | 23+ | PLHCV/HCV- | HIC | Cross-sectional | 348 | 174 | Current use | 53 | 9 |
| Quesada, 2015 | 1994 - 2005 | Thailand | Asia | Community | 18+ | Women | LMICs | Cross-sectional | 50 | 50 | Ever | 14 | 9 |
| Quesada, 2015 | 1994 - 2005 | Nigeria | Africa | Community | 18+ | Women | LMICs | Cross-sectional | 227 | 227 | Ever | 7 | 9 |
| Reed, 2013 | 2003-2006 | USA | North America | Clinic | 20.0 | PLHCV | HICs | Cohort | 289 | 289 | One year | 229 | 8 |
| Roed, 2014 | 2010 - 2011 | Denmark | Europe | Clinic | 51.0 | PLHCV | HICs | Cross-sectional | 120 | 60 | Current use | 32 | 7 |
| Sheehan, 2012 | 2005- 2006 | Argentina | Latin America | Clinic | 18-65 | PLHCV/HIV | LMICs | Cross-sectional | 205 | 61 | Current use | 50 | 7 |
| Shin, 2009 | 2002 - 2008 | Korea | Asia | Clinic | 30+ | PLHCV | HICs | Cross-sectional | 95 | 95 | Current use | 34 | 9 |
| Shuter, 2017 | 2014 - 2016 | USA | North America | Clinic | 51.6 | PLHCV | HICs | Cross-sectional | 77 | 40 | Current use | 5 | 6 |
| Simon, 2014 | 2008 - 2009 | Brazil | Latin America | Clinic | 18+ | PLHIV | LMICs | Cross-sectional | 580 | 138 | Smoking history | 85 | 8 |
| Sims, 2017 | 2014 - 2015 | USA | North America | Clinic | 56.3 | PLHCV | HICs | Cohort | 747 | 747 | Active smoking | 290 | 8 |
| Vahdani, 2009 | Apr - July, 2007 | Iran | Middle East | Clinic | 45±17.7 | Homeless | LMICs | Cross-sectional | 202 | 87 | Smoking history | 66 | 6 |
| Vigani, 2014 | 2009 -2012 | Brazil | Latin America | Clinic | 47(median) | HCV+/ HBV+ | LMICs | Cross sectional | 756 | 348 | Current use | 88 | 8 |
| Weinstein, 2011 | 2001 - 2010 | USA | AMRO | Clinic | 18+ | CLD | HIC | Cross-sectional | 878 | 504 | Smoking History | 154 | 10 |
| White, 2011 | 2007 | USA | AMRO | Clinic | 55.7±6.9 | PLHCV | HIC | Cross-sectional | 91 | 91 | Current use | 33 | 9 |
| Wong, 2015 | 2008 - 2009 | Canada | North America | Community | 18+ | MSM | HICs | Cross-sectional | 1132 | 56 | Current use | 45 | 8 |
| Xu, 2015 | 2012 | China | Asia | Community | 18+ | General population | LMICs | Cross-sectional | 3219 | 1355 | Current use | 838 | 8 |
| Yamini, 2011 | 1998 - 2007 | USA | North America | Clinic | 48.5 ± 10.2 | PLHCV | HICs | Cross-sectional | 800 | 800 | Current use | 462 | 7 |
| Younossi, 2013 | 1999 - 2010 | USA | North America | Community | 18+ | PLHCV | HICs | Cross-sectional | 173 | 173 | Current use | 132 | 9 |
| Zani, 2009 | 2006 - 2007 | Italy | Europe | Clinic | 51.8±11.9 | PLHCV | HICs | Cross-sectional | 229 | 229 | Current use (1cig/day) | 80 | 6 |
| Ziada, 2016 | 2013 - 2015 | Egypt | Africa | Clinic | 55.1 ± 9.8 | PLHCV | LMICs | Cross-sectional | 514 | 514 | Smoking history | 224 | 8 |
